# Optimal timing of booster doses in a highly vaccinated population with minimal natural exposure to COVID-19

**DOI:** 10.1101/2024.05.14.24307386

**Authors:** Eamon Conway, Camelia Walker, Michael Lydeamore, Nick Golding, Gerard Ryan, Dario Mavec, James Oates, Greg Kabashima, David J Price, Freya Shearer, Deborah Cromer, Miles P Davenport, James McCaw, Emily M Eriksson, Philip D Hodgkin, Logan Wu, Thao P. Le, Christopher M. Baker, Ivo Mueller, Jodie McVernon

## Abstract

Population-level waning of protection following immunising exposures is an important determinant of susceptibility to COVID-19 outbreaks. This work outlines an individual-based model (IBM) for the transmission and clinical impact of SARS-CoV-2 that explicitly represents the immunological response to vaccination and infection of each individual. The IBM evaluates waning of immunity to inform risk of infection and related clinical outcomes across a large freely mixing population over time by age and prior exposure status. Modelling immunological responses allows us to investigate the likely impact of immune escape variants based on the landscape in which they emerge. The model described in this paper was motivated by the need to anticipate health and societal impacts of COVID-19 in Australia following emergence of the Omicron variant, in the context of high national vaccine uptake but low infection exposure. It provides a flexible framework for modelling policy-relevant scenarios to inform preparedness and response actions as immunity in a population changes through time.

## 1 Introduction

Understanding population-level immunity against COVID-19 is important for proportionate public health and policy decision making about the imposition of public health and social measures, and vaccine prioritisation and timing, to prevent future surges of infection and disease. Whilst vaccination and prior exposure provide some measure of sustained protection against the clinical consequences of SARS-CoV-2, it is now well established that immunity against infection acquisition wanes over several months [1, 2]. Moreover, variants such as those of the Omicron lineage demonstrate ongoing risks associated with selection for immune escape, reducing effective protection gained from prior vaccine and infection exposures.

Neutralising antibody (NAb) titres provide a measure of functional immune responses induced by SARS-CoV-2 vaccination and infection against sequentially emerging variants. An immunological model has been developed to correlate NAb titres with clinical outcome measures from vaccine efficacy studies to define proxies of protection against a range of infection and disease endpoints [3–5]. The model assumes that NAb titres following vaccination or infection exposure are log-normally distributed in the population and decay exponentially over time, resulting in a loss of efficacy.

While it is thought that other immune mechanisms such as T cells and memory B cells may contribute to defence [6], there remains insufficient evidence to incorporate these mechanisms into a quantitative immunological model.

This immunological model was previously embedded within an existing individual-based model framework that assumed static immune responses [7, 8]. The model presented in this paper extends previous work by explicitly tracking the dynamics of each individual’s NAb titre following vaccination. In this way, it enables exploration of likely transmission and related clinical outcomes of SARS-CoV-2 variants under different vaccination and epidemic scenarios over time. The work was initiated in late-2021, motivated by the need to investigate population-level consequences for Australia of waning vaccine immunity and emergence of immune escape variants, lowering effective protection against COVID-19.

As of early December 2021, Australia had experienced limited and localised SARS-CoV-2 transmission, due to strict inter- and intra-national migration controls and a proactive strong suppression policy. Only 211,000 cases (less than 1% of the population) had been reported nationally, meaning that the vast majority of immunity was derived from vaccination alone, with completed primary course (two dose) coverage of approximately 84% of the eligible population. The country was well advanced on a national reopening plan based on vaccine coverage thresholds deemed sufficient to mitigate impact of the Delta variant predominant at that time [8]. By using a flexible modelling framework that incorporated both immune waning and evasion, we were able to address policy relevant questions about the likely impacts of SARS-CoV-2 Delta and Omicron variants in the early months of 2022 given existing vaccine uptake, and optimal timing of booster vaccine administration to mitigate disease burden.

## 2 Methods

The model framework is comprised of multiple interleaving elements to determine the immune status over time of a modelled population of approximately 8 million individuals, representing a large Australian jurisdiction. Figure 1 provides a schematic of the model framework. First, a data-driven model representing logistics of the Australian vaccination roll-out was used to simulate real world vaccine delivery, including the age-dependent prioritisation over time. Second, viruses with the properties of SARS-CoV-2 Delta and Omicron variants were introduced into the immunised population, and their transmission simulated over time. Each individual’s immune status was tracked following vaccine or SARS-CoV-2 exposure. This immune status influenced whether each new infection exposure would lead to infection acquisition. Finally, a line-list of infections is output from the dynamic transmission model and used as input to a model that captured each individual’s likely clinical pathway based on their age and immune status. By categorising these consequences in relation to distinct clinical presentations we were able to anticipate the likely impact of COVID-19 on the healthcare system. Both the vaccine and clinical pathways models accounted for capacity constraints in health service delivery, allowing simulation of feasible scenarios relevant to public health and policy decision making in context.

**Fig 1.**
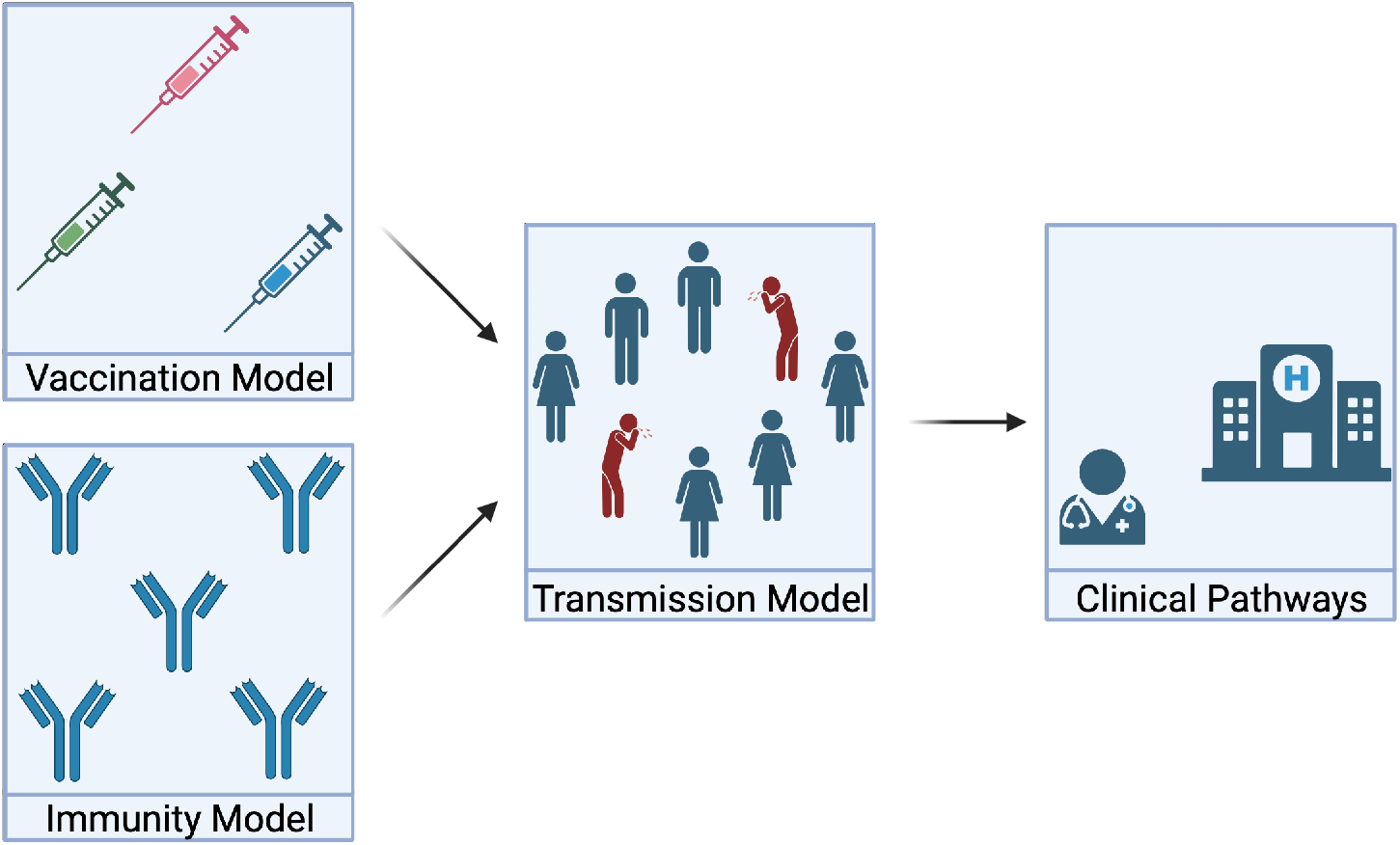
Schematic demonstrating how each component of the model is linked together. First the vaccination allocation is determined and the immunity model is calibrated to real world data. This information is linked into the transmission model, which simulates the spread of COVID-19. Finally, the infections from the epidemic model are linked to the Clinical Pathways model. Created with BioRender.com

### 2.1 Vaccination Model

The model of vaccine roll-out in the Australian population has been developed in house by the Australian Government [8]. In brief, an individual-based model using location and allocation data on vaccine sites around Australia is used to administer available stocks to the seeking population, within supply and delivery constraints [8]. In earlier work, the model was used to investigate alternative primary course allocation strategies, but on this occasion data recorded in the Australian Immunisation Register informed the population’s vaccination status, including the timing of doses and vaccines received. We then explored future roll-out scenarios that included different time windows to booster dose eligibility following completion of the primary series (6 months versus 3 months) and final achieved uptake (80% versus 60%). The model outputs the vaccine type and time vaccination, including primary and booster doses, for each individual in the Australian population. Note that although this study has focused on scenarios directly relevant to Australia, any user defined vaccine roll-out can be simulated and implemented within this framework.

### 2.2 Transmission model

To model the transmission of COVID-19 throughout the population of interest we extended a previous individual-based model [8] to account for loss of protection over time since vaccination. Waning of immunity is accounted for by explicitly modelling the boosting and decay of each individual’s neutralising antibody titre [3, 4].

Each individual within the simulation is assigned an age and corresponding contact profile within an age-based matrix, a predetermined vaccination schedule (using the model described in Section 2.1), and related decay rate of NAbs. Because the vaccination schedule is set elsewhere, we can quickly and efficiently switch out different vaccination scenarios.

The simulation is initialised at a known date, with each individual assumed to be susceptible and to have a neutralising antibody level consistent with their vaccination schedule. This is done by explicitly simulating their previous vaccinations backwards in time, ensuring appropriate initial conditions for population-level immunity. Once an individual is constructed, they are dynamically stored within a vector.

The spread of infection is modelled by directly simulating contact between infectious and susceptible individuals. Since the initial model development, [8], we have updated our model to include over-dispersion in the number of contacts an individual makes.Therefore, for an infectious individual *i* in age bracket *n*, we sample the number of contacts that they make in age bracket *m* from a negative binomial (NB) distribution, such that,

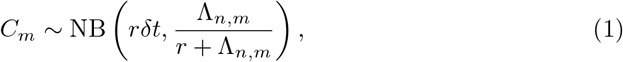

where *C*_*m*_ is the number of contacts made this timestep by the individual in age bracket *m* (in units of contacts per day), *r* is the dispersion parameter, *δt* (day) is the size of the current time step and Λ_*n,m*_ is the mean number of daily contacts between age bracket *n* and *m*. The matrix form of Λ_*n,m*_ used in this work is provided in Section 6.1. Note that we have used the definition of the negative binomial distribution where NB(*r, p*) corresponds to probability density function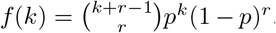. We then sample *C*_*m*_ contacts uniformly from individuals within the *m*-th age bracket. If contact *j* is susceptible we determine if infection occurred using,

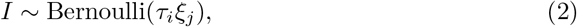

where *I* is an indicator variable for successful infection, *τ*_*i*_ (dimensionless) is the infectiousness of the infectious individual *i* and *ξ*_*j*_ (dimensionless) is the susceptibility of the contact *j*. We note that *τ*_*i*_ is calibrated against a known transmission potential for the population of interest, ensuring that we simulate the appropriate level of disease transmission [8]. The transmission potential encapsulates the intrinsic transmissibility of the circulating virus and the public health and social measures being observed within the population. We also note that both *τ*_*i*_ and *ξ*_*j*_ depend upon the underlying immunological model, which is described in Section 2.4, and the heterogeneous characteristics of the individual of interest (for example, *τ*_*i*_ will be altered depending on the symptom status of the infectious individual and both *τ*_*i*_ and *ξ*_*j*_ will depend upon age). This process of generating contacts and the resulting infections is repeated over all age brackets.

At the point of infection, whether an individual will be asymptomatic or symptomatic is sampled from,

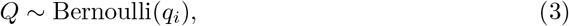

where *Q* is an indicator variable for symptomatic infection and *q*_*i*_ is the probability that individual *i* will be symptomatic, which depends on the age and neutralising antibody titre of individual *i*. We also sample the time that the newly exposed individual becomes infectious, the time for the onset of symptoms and the time that the individual will recover. Given that isolation of COVID-19 cases was mandated in Australia at the time of study, we further simulate the time of isolation in relation to symptom onset, which reduces the individual’s duration of infectiousness. Note that each of these times are sampled from known distributions based on local case data [9]. These data have also been used to estimate the likely constraining impact on transmission of active case and contact finding and management, collectively termed test, trace, isolate, quarantine (TTIQ). We include a factor to account for a ‘partial TTIQ’ effect under high case loads as estimated from local data in earlier work [8].

When an infectious individual recovers, we store the individual’s age, time of symptom onset, neutralising antibody titre at exposure, symptom status (*Q*), the number of individuals they infected, their vaccine status (what was the latest vaccine they received) at exposure, the time they were isolated from the community and the number of times that they have been infected. This generates a line-list of infections that is used to model clinical outcomes (Section 2.3).

### 2.3 Clinical outcomes model

The clinical outcomes model is based on and extends the previously described clinical model [7, 8, 10]. This implementation extends on previous work by restructuring the model as a continuous-time, stochastic, IBM, where the NAb titre and age of the individual determines their transition probabilities. The full compartmental structure of the clinical outcomes model is depicted in Figure 2.

**Fig 2.**
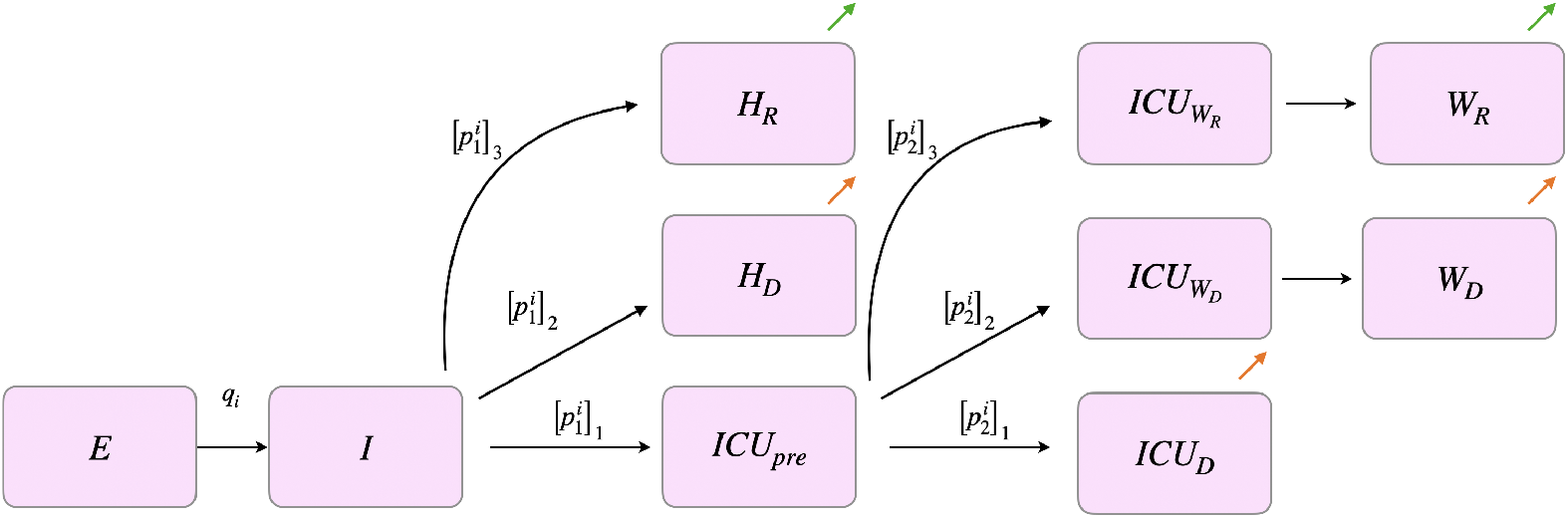
Diagram representing the transitions an individual can make in the clinical pathways model. Transitions resulting in discharge and death are represented with green and orange arrows.

All parameters governing the pathway each individual takes through the health system are altered depending upon their individual NAb titre. The evaluation of these transition probabilities are explained in full in Section 2.4 and baseline parameters associated with Delta severity in unvaccinated individuals are given in Section 6.6.

For symptomatic individual *i* in the line-list output from the transmission model, we determine if they are hospitalised by sampling,

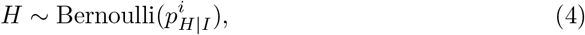

where *H* is an indicator variable for hospitalisation and 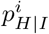 is the probability that individual *i* is hospitalised given symptomatic infection.

There are three initial pathways for hospitalised individual *i*. Individual *i* will either recover and be discharged from a ward bed, die in a ward bed, or move to an ICU bed; as the three pathways have different length of stay distributions they are modelled as three separate compartments *H*_*R*_, *H*_*D*_ and ICU_*pre*_. To determine which pathway individual *i* will follow, we sample from,

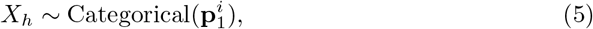

where *X*_*h*_ is the sampled hospital pathway,

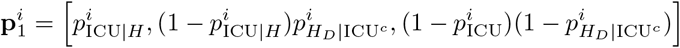

is a vector containing the probability of transitioning into ICU_*pre*_, *H*_*D*_, or *H*_*R*_ respectively, 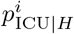 is the probability that individual *i* is admitted to ICU given they are hospitalised and 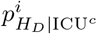 is the probability that individual *i* dies on ward given that they are in hospital and are not going to ICU. If individual *i* requires the ICU, they follow a further ICU pathway to determine their final outcome.

There are three possible pathways for an individual in the ICU. The individual will either die in the ICU (ICU_*D*_), die in a ward bed after leaving the ICU (ICU_*WD*_), or recover and be discharged from a ward bed after leaving the ICU (ICU_*WR*_). We sample which pathway is taken within the ICU from,

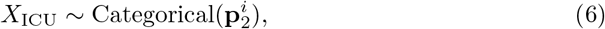

where *X*_ICU_ is the sampled ICU pathway,

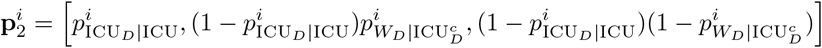

which is a vector containing the probability of transitioning into the ICU_*D*_, ICU_*WD*_ or ICU_*WR*_ compartments respectively, 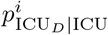 is the probability that individual *i* dies in the ICU given they were admitted to ICU and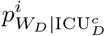 is the probability that individual *i* dies in a ward bed after leaving ICU without dying. For an individual that transitions into ICU_*WR*_ or ICU_*WD*_, they will move into a post-ICU ward compartment, *W*_*R*_ or *W*_*D*_, where they will either recover or die respectively.

Finally, the length of stay for individual *i* in each compartment is sampled such that,

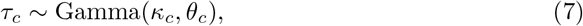

where *τ*_*c*_ is the time spent in compartment *c*, and *κ*_*c*_ and *θ*_*c*_ are the shape and rate parameter for compartment *c* respectively. Uncertainty is incorporated by sampling rate and shape parameters from the posterior estimated for the Australian Delta wave [11]. The mean lengths of stay in each compartment by age are given in Supplementary Table 3. Note that we assume that neutralising antibody titre levels do not change the distribution of time spent in any compartment.

By generating a clinical timeline for every symptomatic individual, we can calculate hospital admissions, ICU occupancy, ward occupancy and deaths by age in continuous-time. Furthermore, by explicitly incorporating the effects of neutralising antibodies on protection against each outcome, we are able to account for individual level immune responses.

### 2.4 Immunological model

Within both the transmission model (Section 2.2) and the clinical outcomes model (Section 2.3) we include an immunological response to COVID-19. This immunological response is handled by directly modelling each individual’s neutralising antibody (NAb) titre. By using the model of [3] and [4] we can relate an individual’s NAb titre to their protection against all outcomes of interest: infection, symptomatic disease, onward transmission given breakthrough infection, hospitalisation and death. The fact that each individual is assigned their own NAb titre results in inter-individual variation that leads to varying distributions of protection throughout the community, subsequently impacting the transmission dynamics. In the Australian context, by the time Delta and Omicron outbreaks occurred almost all immunity was vaccine-derived, therefore the model was initialised by making the plausible simplifying assumption that NAb titres were derived from the vaccine roll out alone.

An individual’s NAb titre can be increased by a variety of exposures. The processes that we consider are: the first, second and booster dose of a vaccine, or infection occurring in an unvaccinated or vaccinated individual. Note that for simplicity we assume that infection prior to or following vaccination results in the same titre of NAb. Immune responses are stratified by the type of vaccine product, AstraZeneca (AZ) or mRNA vaccine (Pfizer or Moderna), that the individual has received based on the model from Section 2.1.

At the time of a boosting process, the level of NAb titre that is acquired is sampled from,

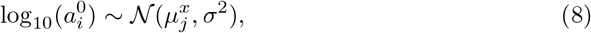

where 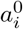 is the NAb titre that individual *i* is boosted to, 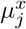 is the mean NAb titre against strain *x* of the population after boosting process *j* and *σ*^2^ is the variance of NAbs across the population at the time of boosting.

The mean NAb titre, 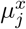, is specified using the following rules based upon the infection and vaccination history of the individual. In this work it is assumed that an unvaccinated individual will have an average NAb titre of 0.0 on the log_10_-scale after exposure. This is our baseline measurement and is used to calibrate across multiple neutralising antibody studies. For an individual that has no prior exposure to COVID-19, their vaccination induces an antibody response such that,

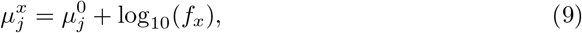

where 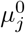 is the mean level of NAb titre for vaccine *j* against a base strain of COVID-19 (for us this is Delta) and *f*_*x*_ is the fold change in NAb titre between the base strain and strain *x*. To account for the effect of exposure to COVID-19 prior or post vaccination, we use an altered form of Equation 9. For brevity, we have used Table 1 to list the equations used to obtain 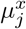 with infection.

**Table 1.** Exposed individuals: Assumed relationships within the immunological model for NAb titre for individuals that have been exposed to COVID-19. The extended subscript with an E represents prior or current exposure to the circulating strain of COVID-19.

| Processes | Average titre formula |
| --- | --- |
| Unvaccinated (U∩E) | $\mu_{U\cap E}^x = \mu_U^0$ |
| AZ dose 1 (AZ1∩E) | $\mu_{AZ1\cap E}^x = \mu_{P2}^0$ |
| AZ dose 2 (AZ2∩E) | $\mu_{AZ2\cap E}^x = \mu_B^0$ |
| Pfizer dose 1 (P1∩E) | $\mu_{P1\cap E}^x = \mu_{P2}^0$ |
| Pfizer dose 2 (P2∩E) | $\mu_{P2\cap E}^x = \mu_B^0$ |
| mRNA booster (B∩E) | $\mu_{B\cap E}^x = \mu_B^0$ |

It is assumed that an individual’s titre of neutralising antibodies will decay after boosting. This decay is assumed to be exponential, therefore,

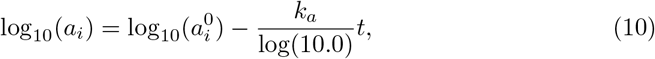

where *a*_*i*_ is the time dependent NAb titre of individual *i, k*_*a*_ is the decay rate of neutralising antibodies and *t* is the time from the last boosting process. In this work we have assumed that *k*_*a*_ is constant for all time.

An individual’s protection against disease outcome *α*, denoted by *ρ*_*α*_, is a function of their current antibody titre. The functional form of *ρ*_*α*_ is assumed to be logistic and is governed by,

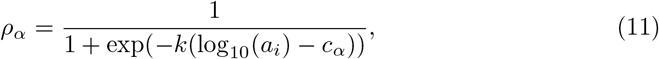

where *k* is governs the steepness of the logistic curve (logistic growth rate), and *c*_*α*_ defines the midpoint of the logistic function for disease outcome *α*.

The immunological model interacts with the transmission model by altering the probability that an individual develops symptoms, *q*_*i*_, their rate of onward transmission given breakthrough infection, *τ*_*i*_, and the contact’s level of susceptibility, *ξ*_*j*_.

The susceptibility of contact *j* is,

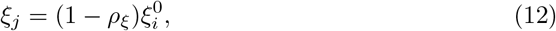

where *ρ*_*ξ*_ is the protection against infection and 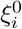 is the susceptibility of the *i*th individual if they were completely COVID naive. The probability that individual *i* develops symptoms is governed by,

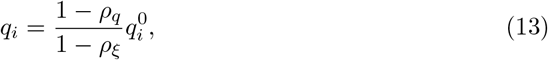

where *ρ*_*q*_ is the protection against symptomatic infection and 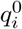 is the probability of symptomatic infection for individual *i* if they were completely COVID naïve, i.e., have zero NAb titre.

The expressions for onward transmission rate are more complex. It is assumed that asymptomatic individuals are 50% less likely to infect their contacts when compared to their symptomatic counterpart [8]. However, this reduction in transmission due to asymptomatic infections is not accounted for in the clinical trial data used to calibrate the protection against onward transmission. To avoid double counting the effect of the NAbs we alter the functional form for the rate of onward transmission to,

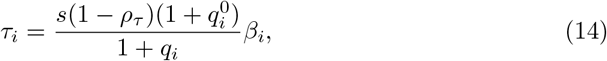

where *s* is either 0.5 or 1 depending upon whether the individual is asymptomatic or symptomatic respectively, *ρ*_*τ*_ is the protection against onward transmission and *β*_*i*_ is the baseline (i.e., zero NAb titre) infectiousness of the infector. For a full derivation of Equation 13 and Equation 14, see Section 6.3.

The clinical outcomes model uses the transmission model as an intermediary between the immunological response of each infected individual and their corresponding clinical outcome. This is done by outputting each infected individual’s NAb titre at the point of exposure, a symptom indicator and their time of symptom onset for use within the clinical pathways model.

The immunological model determines the probability of hospitalisation, ICU requirement and death based on observed relationships between NAb titres and clinical endpoint outcomes from efficacy studies [3–5]. For a symptomatic individual *i*, the probability of hospitalisation is given by,

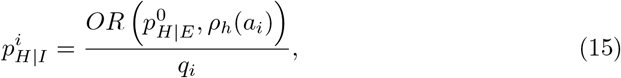

where 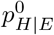 is the baseline probability of hospitalisation given infection, *ρ*_*h*_(*a*_*i*_) is the protection against hospitalisation, *a*_*i*_ is individual *i*’s NAb titre at the point of exposure, and,

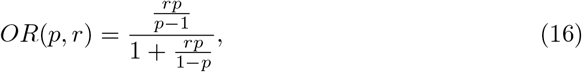

is the function that uses odds ratio *r* and baseline probability *p* to compute an adjusted probability.

If individual *i* is hospitalised, the probabilities governing which hospital pathway is chosen are altered such that,

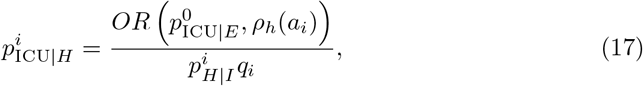

and,

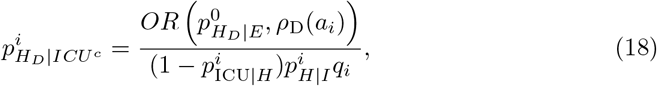

where 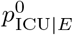 is the baseline probability of requiring the ICU given infection, 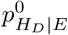 is the probability of death on ward (without visiting ICU) given infection and *ρ*_D_(*a*_*i*_) is the protection against death given infection.

If individual *i* is in the ICU, then their probabilities of death in the ICU, 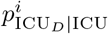, and death on the ward given they left ICU without dying, 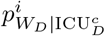, are altered such that,

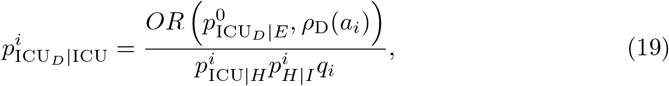

and,

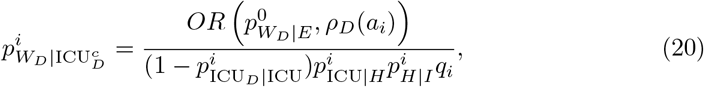

where 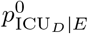 is the baseline probability of dying in the ICU and 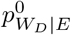 is the baseline probability of dying in the ward after returning from the ICU. Note that we assume no difference between the protection from hospitalisation given infection and the protection from ICU given infection here.

To determine all parameters in Equation 10 and Equation 11, we use a re-implementation of [3] and [4] in a Bayesian framework [12]. This allows us to calibrate the level of protection, which is analogous to vaccine efficacy for individuals with no exposure to COVID-19, to real-world vaccine effectiveness studies. The model fit in [12] takes in a range of data relating NAb levels to efficacy, and estimates of vaccine effectiveness from a range of studies to estimate effectiveness over time against the Delta variant. To estimate the effectiveness against the Omicron variant, Golding and colleagues estimate an ‘escape’ parameter for the Omicron variant relative to the Delta variant. This was done by using the relative rates of infection in Danish households between Omicron and Delta to estimate the the relative *R*_0_ between the variants [13], and early evidence of vaccine effectiveness against Omicron from the UK to understand the level of vaccine escape [14]. This was then combined with the information fit on the Delta variant to model waning over time for both the Delta and Omicron variants.

Figures 3 and 4 show the levels of protection against all outcomes of interest for both the Delta and Omicron strain, using parameters in Table 2. Notably, the lowest protection results from vaccination with AZ, which was the initially recommended vaccine for older individuals in Australia.

**Table 2.** Estimated parameter values from the immunological model.

| Parameter | Value | Parameter | Value |
| --- | --- | --- | --- |
| $c_\tau$ | 0.02953683449343693 | $\log_{10}(f_{\text{Delta}})$ | 0.0 |
| $c_\xi$ | -0.47195962651907175 | $\log_{10}(f_{\text{Omicron}})$ | -0.6923808174384031 |
| $c_q$ | -0.6442020773907903 | $\mu_U^0$ | 0.0 |
| $c_h$ | -1.2161786725147814 | $\mu_{\text{AZ1}}^0$ | -0.5299522095755013 |
| $c_d$ | -1.1753151405677293 | $\mu_{\text{AZ2}}^0$ | -0.12031713312180076 |
| $\log(k)$ | 1.686059432639791 | $\mu_{\text{P1}}^0$ | -0.23154132545543396 |
| $\sigma$ | 0.4647092 | $\mu_{\text{P2}}^0$ | 0.1540166902000597 |
| $k_a$ | 0.008235096361537353 | $\mu_B^0$ | 0.3225538899068383 |

**Fig 3.**
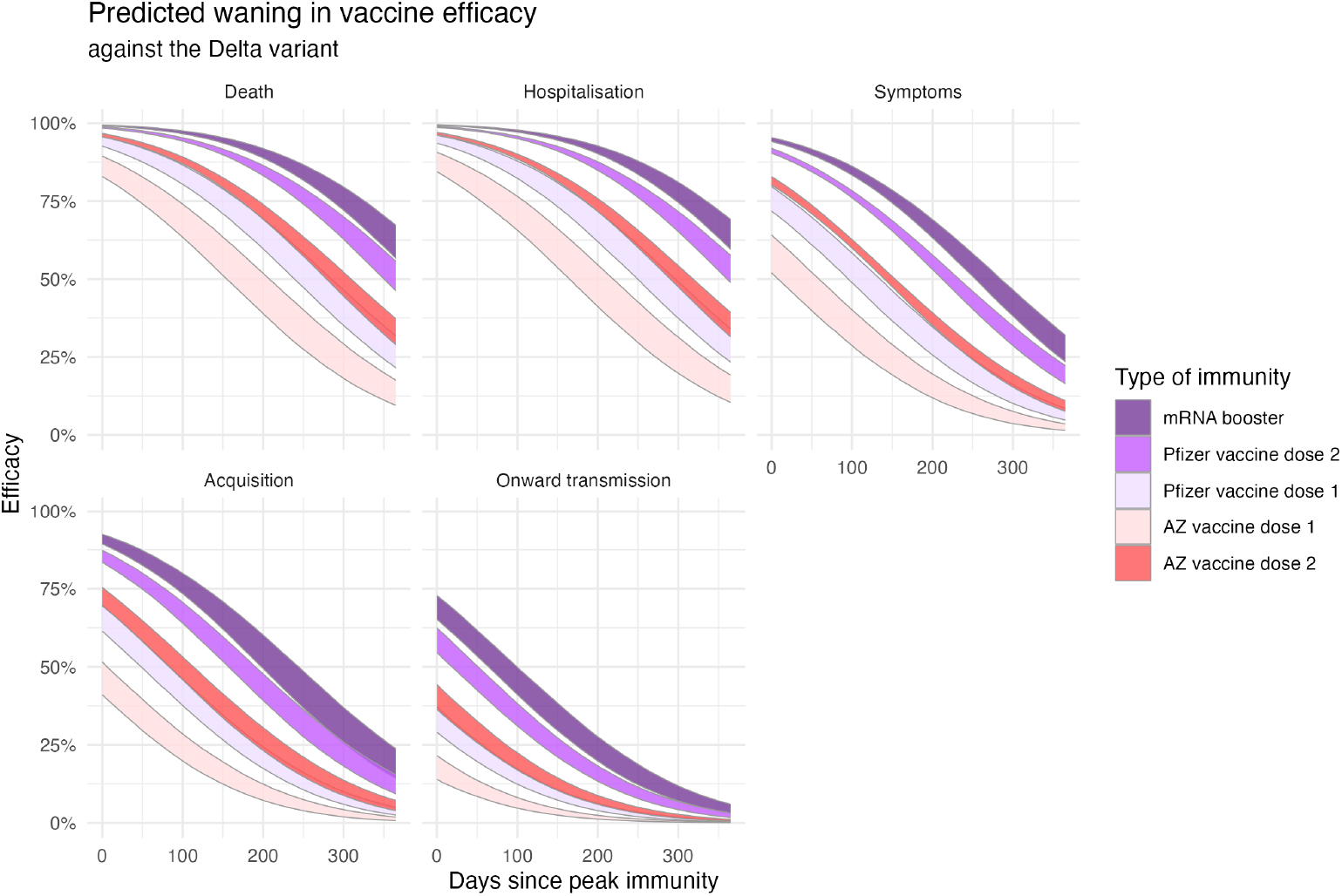
Estimated levels of protection for all outcomes of interest against the Delta strain (modified from [12]). The bands correspond to the 90% confidence interval.

**Fig 4.**
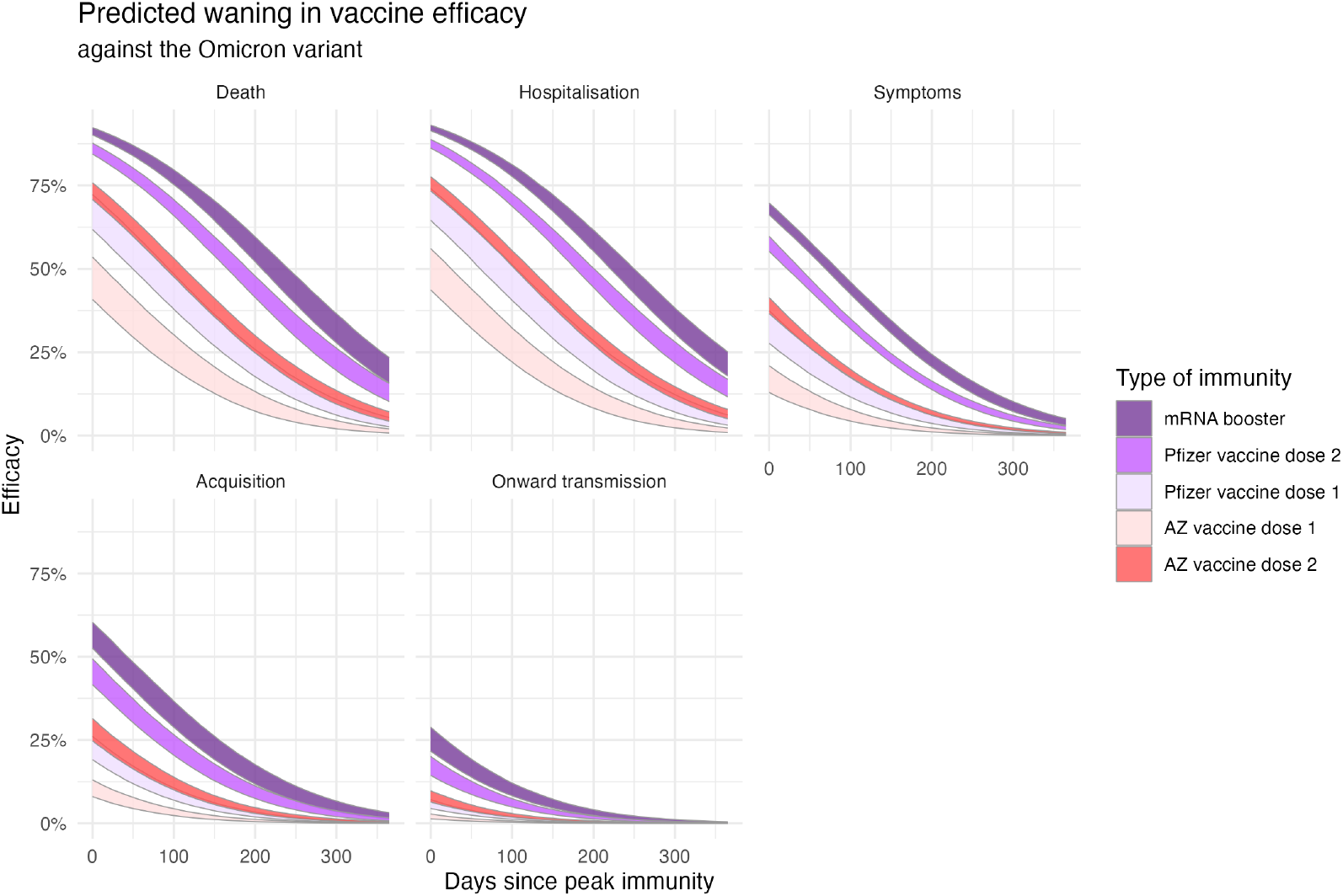
Estimated levels of protection for all outcomes of interest against the Omicron strain (modified from [12]). The bands correspond to the 90% confidence interval.

## 3 Results

### 3.1 Vaccination Scenarios

The initial immune status of the population was determined by the data-driven model of the Australian COVID-19 vaccine roll-out, as described in Section 2.1. The vaccine program was initially targeted at older age groups who were at higher risk of severe outcomes, with later phases opened to peak transmitting populations with a focus on reducing transmission towards reopening goals [8]. Presented in Figure 5 are the four future booster implementation scenarios determined in consultation with the Australian Department of Health. These were chosen to assess the impact of reducing time between completion of the primary series and booster dose administration — from the then recommended six months, to three months — with either 60% or 80% final uptake achieved in eligible cohorts. Given the age staggered implementation of the initial vaccine program, eligibility for the booster dose followed the same staggered order of administration, meaning that individuals at highest risk of disease were immunised first in all scenarios.

**Fig 5.**
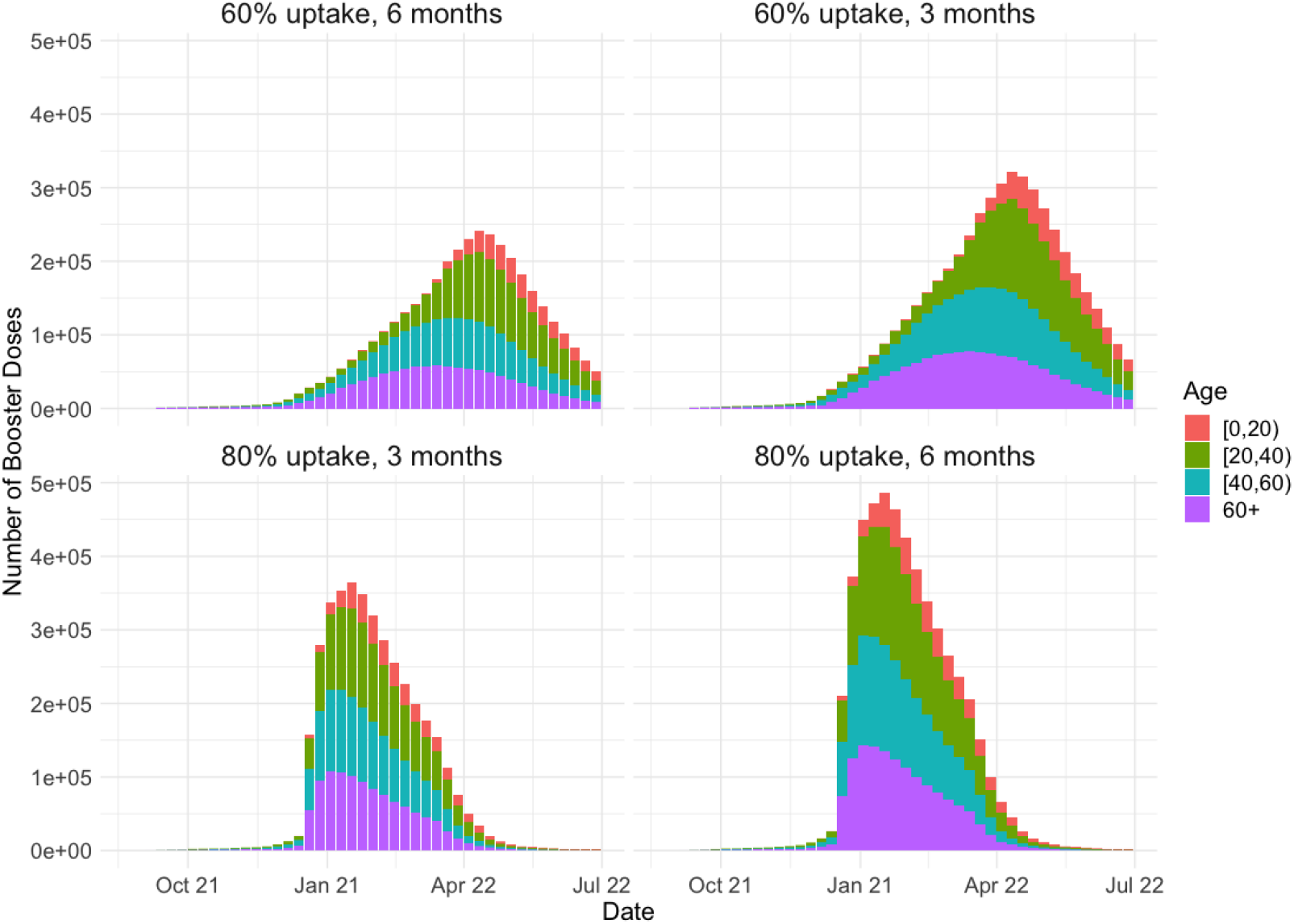
Daily number of booster doses administered in four representative age groups. Due to the age structured administration of the primary course, we can see that younger individuals receive their booster doses after older individuals.

### 3.2 Waning immunity and booster requirement

We hypothesised that due to the waning of immunity, the Australian population was prone to resurgence of COVID-19 infections even if Delta remained the predominant circulating variant. To investigate the possible impact of waning immunity through the national reopening phase, we seeded 1,000 exposures of the Delta variant on 22nd November 2021, reflecting known circulation in Australia’s Eastern states at that time. Finally, we set the transmission potential to 6.32, corresponding to a reproduction number estimated for the Delta variant in the Australian context under minimal public health and social measures and assuming no impact of TTIQ on transmission [8].

Our model anticipates that loss of primary course protection over early 2022 would drive an early autumn resurgence of infections Figure 6. This wave could be ameliorated to some extent by boosting with a 6 month delay, but strongly suppressed by reducing the interval between second and third doses to 3 months. Similar benefits of accelerating the booster program are observed in Figure 7 when achieved booster uptake is only 60%, however the peak number of daily cases exceeds those for the 80% coverage case.

**Fig 6.**
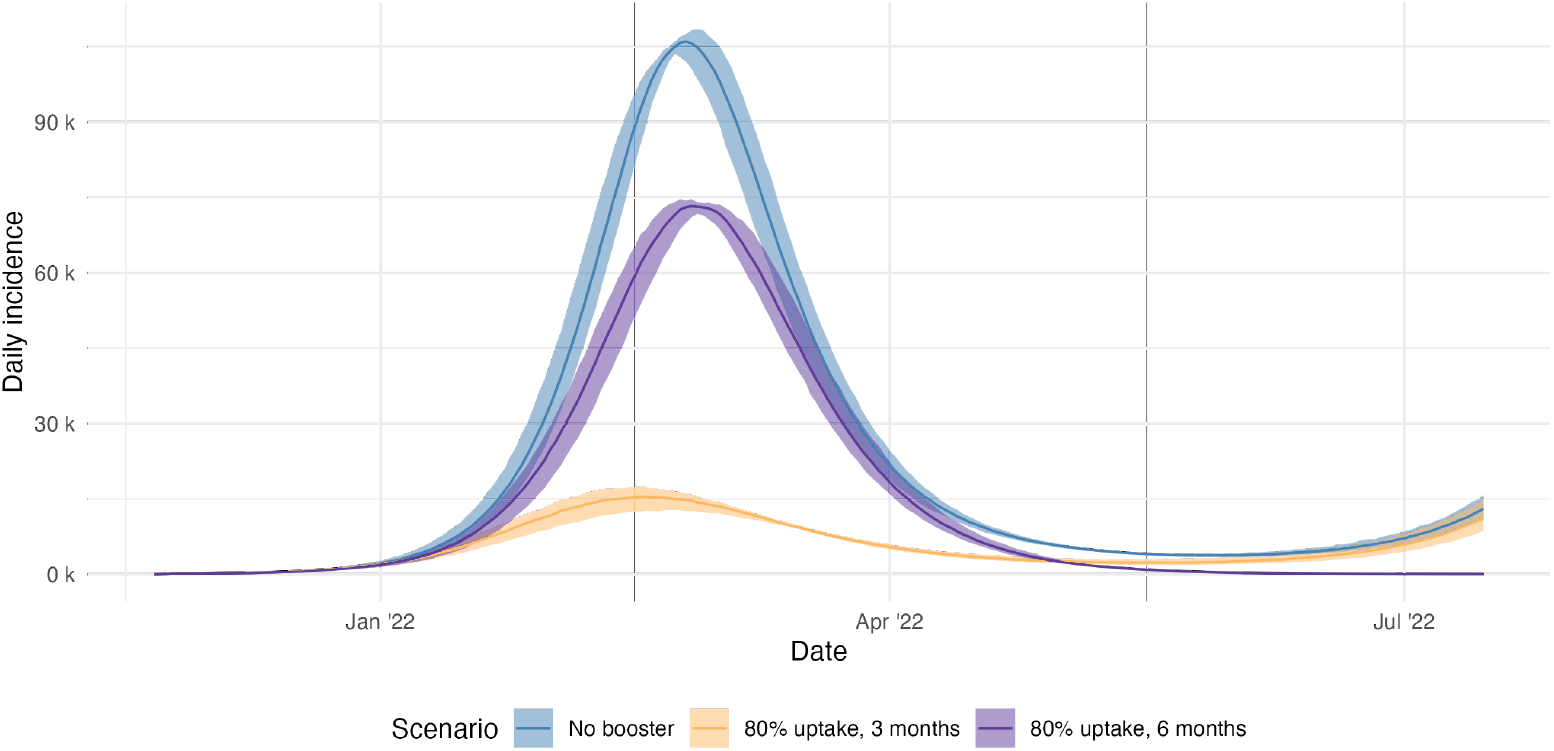
Daily incidence of all infections for the Delta variant only under alternative immunisation scenarios. We compare no boosting as a baseline with 80% of the primed (vaccinated) population for either the (initial) 6 or (proposed) 3 month delay to booster eligibility.

**Fig 7.**
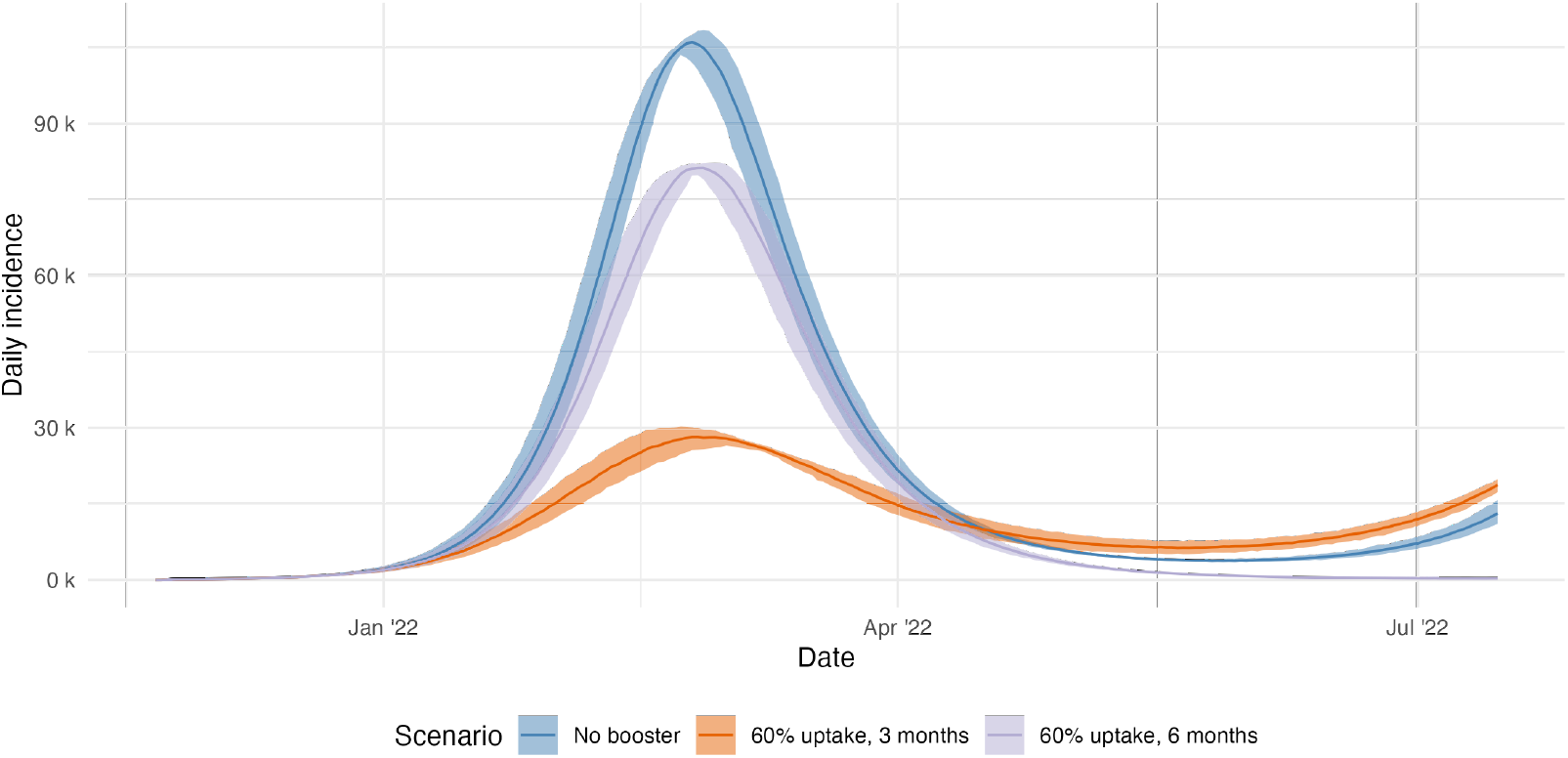
Direct comparison of the simulated daily incidence of infections for the scenarios with no boosters, a 3 month interval at 60% coverage, and a 6 month interval at 60% coverage.

The projected impacts of the Delta variant on the health care system are shown in Figure 8. As would be anticipated from the corresponding infection curves (Figure 6), both hospitalisations and deaths are decreased as the booster interval shortens.

**Fig 8.**
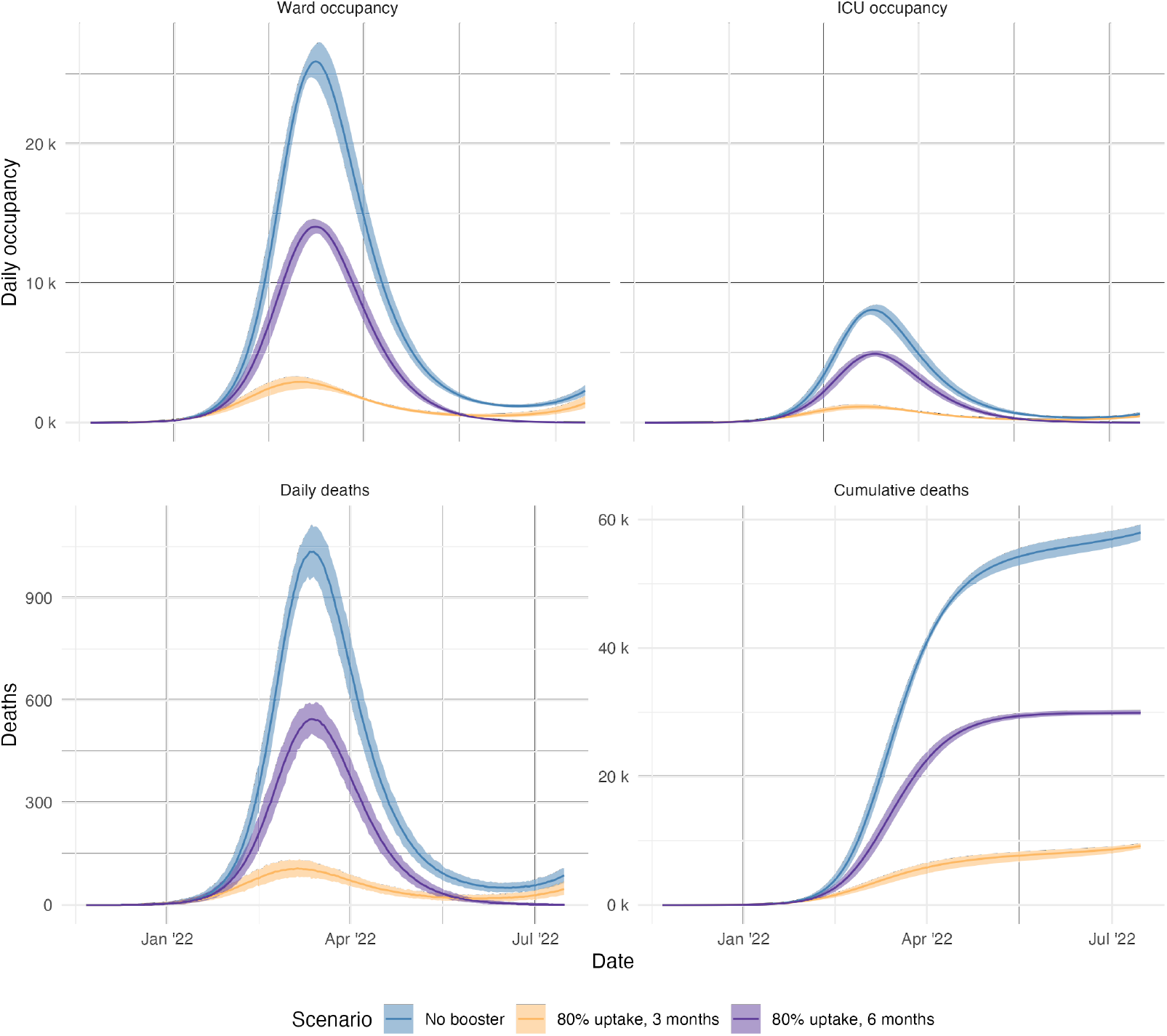
Simulated Ward and ICU occupancy as well as the daily and cumulative deaths for each Delta scenario.

Substantive health benefits are still observed if booster uptake in the eligible populations reaches only 60% (see Supplementary Figure 11, with timeliness being a more critical determinant of outcomes than coverage.

### 3.3 Emergence of Omicron variant

Due to the reported immune escape and transmissibility of the Omicron variant in late 2021, we modelled the potential impacts of an Omicron outbreak in Australia. Using the model of [12], we estimated a 4.924-fold reduction 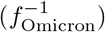 in neutralizing antibodies and an increase in intrinsic transmissibility of the Omicron variant of 1.113 times compared to the Delta variant. Importation of the variant through international arrivals was accounted for by seeding 30 exposures on 22 November 2021 into the modelled population. The population’s immunity status was configured according to the same vaccine priming and boosting scenarios explored for the Delta variant Section 3.

For all scenarios considered, Omicron spreads rapidly throughout the population (see Figure 9), resulting in a peak incidence of daily infections far beyond those anticipated for Delta. For these results, we assume 80% booster vaccination uptake (for 6 and 3 month eligibility intervals) compared with the base case of no booster program. This finding reflects the predicted level of immune escape of the Omicron variant, enabling acquisition and onward transmission of infection. Bringing forward the booster program has only marginal impact on epidemic dynamics.

**Fig 9.**
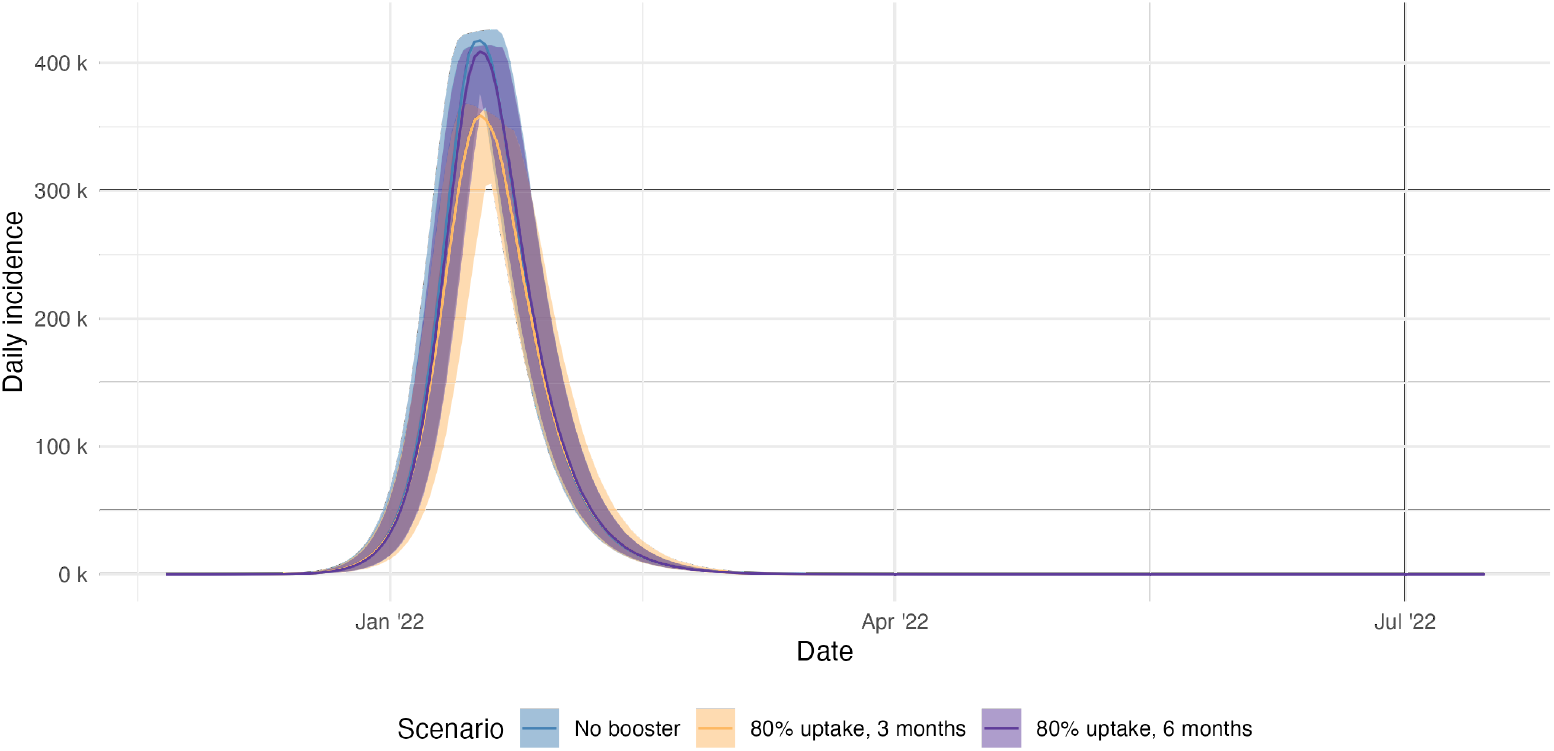
Simulated daily incidence of infections for the Omicron variant only under alternative immunisation scenarios. We compare no boosting as a baseline with 80% of the primed (vaccinated) population for either the 6 or 3 month delay to booster eligibility.

At the time of investigation the likely clinical impact of Omicron in the Australian population was highly uncertain. Initial observations from South Africa [] indicated that the intrinsic severity of Omicron was greatly reduced in a population with a high level of exposure to SARS-CoV-2 across multiple infection waves [], but there was no prior experience of the virus in settings where immunity was principally vaccine-derived. To explore this critical uncertainty in the model we investigated four possible levels of severity for Omicron within the clinical outcomes model for the 80% uptake within three month scenario, the results of which are shown in Figure 10. These levels of severity were explored by multiplying the baseline hospitalisation, ICU and death probabilities *p*_*H*|*E*_, *p*_*ICU*|*E*_, 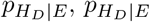, and 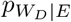 (as defined in 6.6) by various values. The worst case scenario assumed severity equivalent to the Delta variant, ranging down to a best-case of only one tenth that severity, with markedly different impacts for health sector capacity.

**Fig 10.**
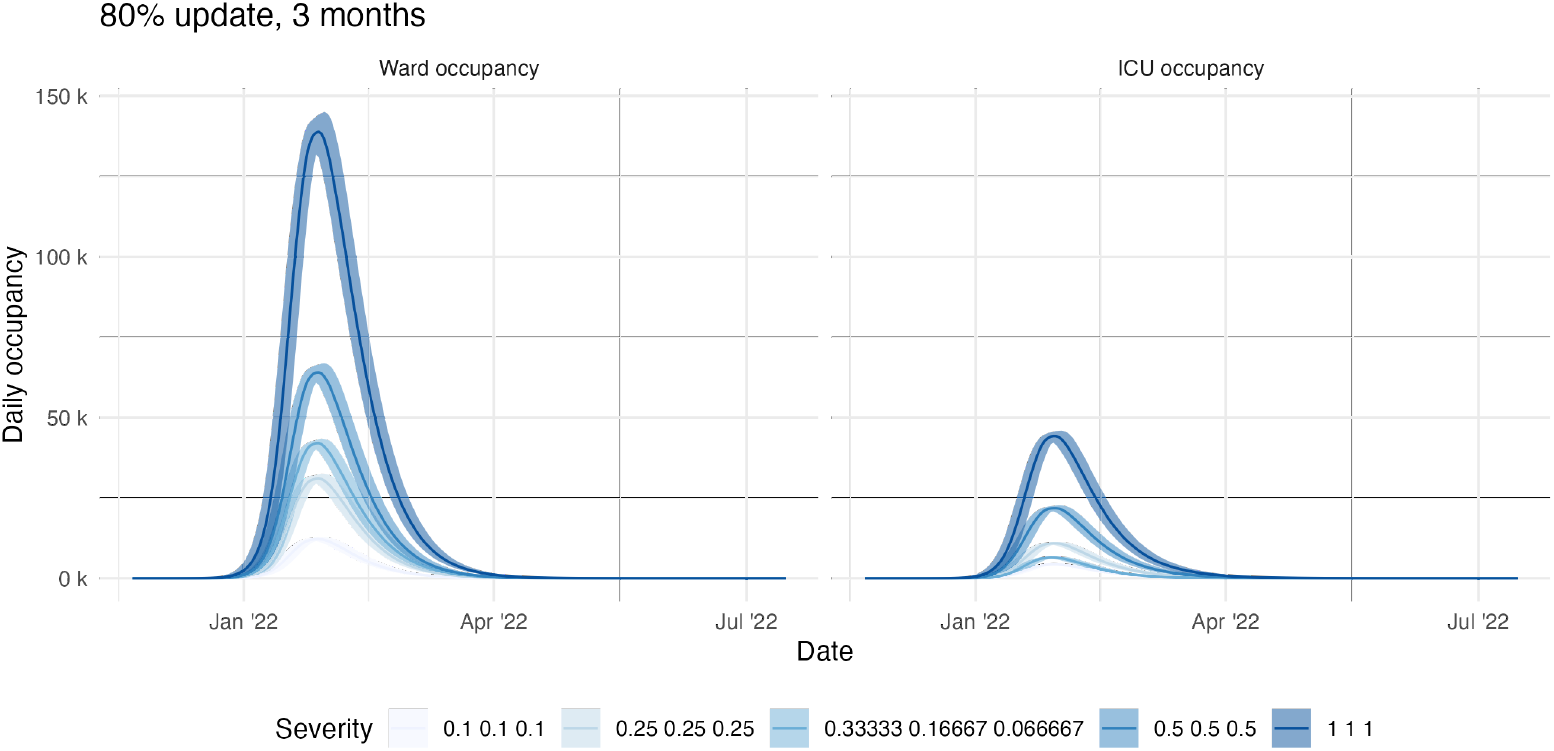
The effect of reduced levels of severity on the clinical outcomes pathways.

## 4 Discussion

This work demonstrates the utility of a flexible model framework to consider the likely impact of emerging SARS-CoV-2 variants of concern in the context of a given population’s immune landscape. In this example, we estimated the impact of the Omicron variant in a highly immunised population and used these results to support policy decision making for the Australian COVID-19 booster vaccination program. The findings presented in this work informed the Australian Government response to accelerate administration of (third) booster doses nationally from late-December 2021.

Due to extremely high vaccination coverage Australia had started implementing its re-opening strategy, with a shifted focus from minimising transmission to the mitigation of severe disease in line with recommendations endorsed by SAGE and WHO [15, 16]. This necessitated the need to focus on control measures that were not strict mobility restrictions and stringent public health and social measures. As such, booster doses were seen as an integral component in the re-opening strategy. Our results indicated that the booster dose eligibility interval should be shortened to three months, maximising the population level of protection against transmission and severe disease. In doing so, the spread of the Delta variant was greatly diminished and the population had high levels of protection against severe clinical outcomes. We note that our results for the daily number of infections indicated that booster doses, irrespective of the booster dose eligibility interval, cannot control the spread of Omicron. This is due to the immune escape component of the Omicron variant. However, by shortening the booster eligibility interval there will be a boost in protection that minimised potential severe clinical outcomes.

The large estimated impact of accelerating the booster roll out on clinical outcomes was partly due to the age-based prioritisation of the primary course of vaccination in Australia. Australia’s initial COVID-19 vaccine roll out was focused on those at most risk of severe disease, including older individuals and the immune compromised.

Consequently, we anticipated substantial waning of vaccine-derived immunity in these groups by the end of 2021. Given that individuals were only eligible for a booster dose after a finite minimum interval from the primary vaccine series, these same high risk groups were once again the first eligible for the third dose. This priority order resulted in the greatest achievable impact on clinical outcomes in the early stages of the roll out, as initial delivery was to those at most risk of severe disease.

A strength of our work is the flexibility of combining multiple interleaving models to accurately characterise the spread of COVID-19 in the population of interest.

Throughout the COVID-19 pandemic, multiple research groups were working on separate mathematical models for projects that were being used to help inform Australian Government policy. This enabled the compartmentalisation of approaches, with each group able to work concurrently. However, this limits the ability of answering broad questions that requires modelling results from across a range of working groups. This work provided a unified framework that incorporated all necessary models to accurately simulate the transmission of COVID-19. Importantly, this framework can quickly adapt to changes in any component of the model, providing the flexibility and speed that is essential to answer policy relevant questions.

To date, there is ongoing uncertainty about the durability of vaccine protection against severe outcomes and optimal dosing intervals. As such it is important to remain alert to the potential of more severe variants arising and spreading rapidly through the population, and to understand which immunisation strategies could minimise the impact of these variants. Due to the flexibility of the immunity model used within our model framework, we are easily able to adapt our approach to such considerations, even applying the model to other countries.

A limitation of the framework presented in this paper is that the epidemic model dynamics are independent from the clinical pathways model and vaccine roll out. This independence allows for greater flexibility, so that other epidemic models for different purposes could feed into the clinical pathways model. The main drawback from this flexible approach is that the clinical pathways model does not update the status of people in the epidemic model: as a result, people can die in the clinical pathways model and continue to be counted as infectious or reinfected in the epidemic model. We confirmed numerically that this double counting did not impact substantively on transmission given the small numbers of deaths relative to the overall population size.

A further limitation of this work is the application of TTIQ. Within our transmission model, we assumed consistent TTIQ capacity over time, irrespective of the total number of infections. In reality, the effectiveness of such system declines as the number of infections increases and exceeds public health capacity. If TTIQ effectiveness were to decline over time, the interval between infection and isolation would increase, resulting in a higher effective reproduction number. Individuals may also change their behaviour when the perceived risk of infection is increased, for example, when case numbers are high. This behaviour change may take the form of voluntary isolation or social distancing. We do not investigate either of these effects in this work.

This work was based on the population of Australia around the time of the first Omicron wave, so only a small proportion of the population had infection-derived immunity. Thus, we initialised the model with a population immunity determined solely by the vaccine roll out. This paper has not presented a method for initialising the model in a context with a more complex history of vaccination and waves of infection. The complex history of vaccination and waves of infection will be a topic of future investigations [17].

## 5 Conclusions

We present an efficient implementation of a model which captures epidemic dynamics at both the individual immunity level and the population-level. This allows the population-level infectious disease dynamics to be governed by immune-history and therefore is appropriate for understanding varied experiences of outbreaks in different locations. For example, in Australia, immunity prior to the Omicron outbreak was almost entirely governed by the vaccine roll out, whereas in many other countries, population-level immunity to BA.1 may have largely been determined by recent large outbreaks of ancestral strains.

This study incorporated a continuous waning model of neutralising antibodies, which allows protection against different end points to change through time. As a result, this study was able to consider both infection and clinical endpoints to answer policy-relevant questions, such as the importance of the timing of booster eligibility. Our study indicates that reducing the time between the primary and booster dose from six months to three months in Australia was an effective way of reducing morbidity and mortality of the Omicron wave. This is because the immunity from the primary dose had waned significantly by the time Omicron emerged, and the three month scenario provided the greatest population-level protection through the first quarter of 2022.

This study shows that although booster doses are a highly effective protection against transmission and clinical outcomes for the Delta variant, vaccines are a less viable strategy against the Omicron variant. Our scenario analyses shows that a hypothetical strain with the immune-evasion characteristics of the Omicron variant and the severity characteristics of the Delta variant would have had catastrophic clinical impacts. Omicron proved to have a greatly reduced severity compared with ancestral strains [18–20], but its high inherent transmissibility and ability to evade immunity still resulted in a substantive burden on public health and clinical systems.

Lastly, this study captures an example of real, rapid, innovative research done by a large interdisciplinary team in response to a public health crisis, providing a record of the methods and results used to advise and public health policy governing tens of millions of people.

## Data Availability

All data produced in the present study are available upon reasonable request to the authors

## 6 Supporting information

### 6.1 Contact Matrix

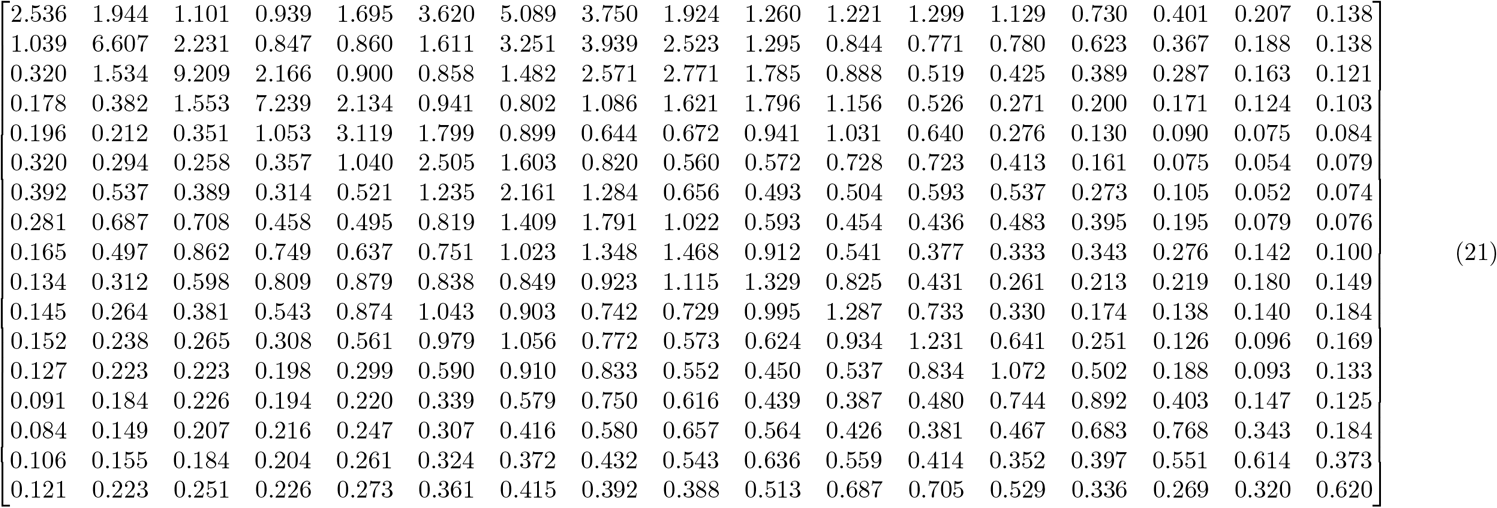

### 6.2 60% uptake results

**Fig 11.**
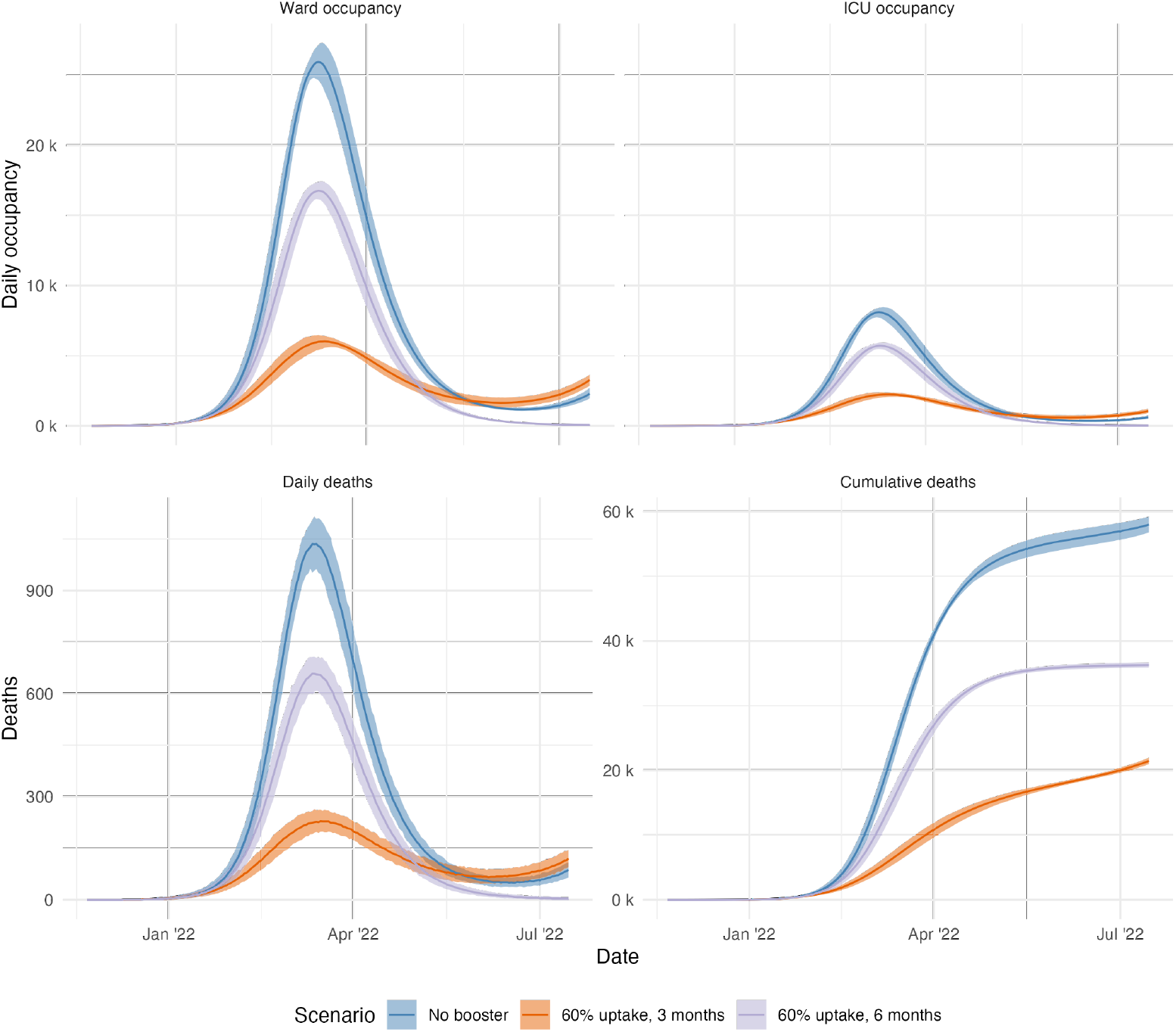
Above: Daily deaths and cumulative deaths for the projected Delta scenarios at 60% uptake. Below: Daily deaths and cumulative deaths for the projected Delta scenarios at 60% uptake.

### 6.3 Sensitivity analysis public health and social measures

### 6.4 Derivation of *q*_*i*_ and *τ*_*i*_

To determine the functional form of *q*_*i*_ and *τ*_*i*_, we need to reconcile the differences between the vaccine efficacy calculated in the immunological model and the conditional protection estimates required within the transmission model.

#### 6.4.1 Protection against acquisition

Protection against acquisition is arguably the simplest to measure using a sero-prevalence survey. It is possible to detect all asymptomatic and symptomatic infections in your cohort using only observations at the start and end of the study. Given the known vaccine status of each individual it is then trivial to calculate the protection against infection. Therefore, we can express the probability of being infected given you are vaccinated as

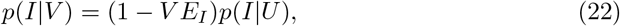

where *V E*_*I*_ is the vaccine efficacy against infection and *p*(*I* |*U*) is the probability of infection given you are unvaccinated.

**Fig 12.**
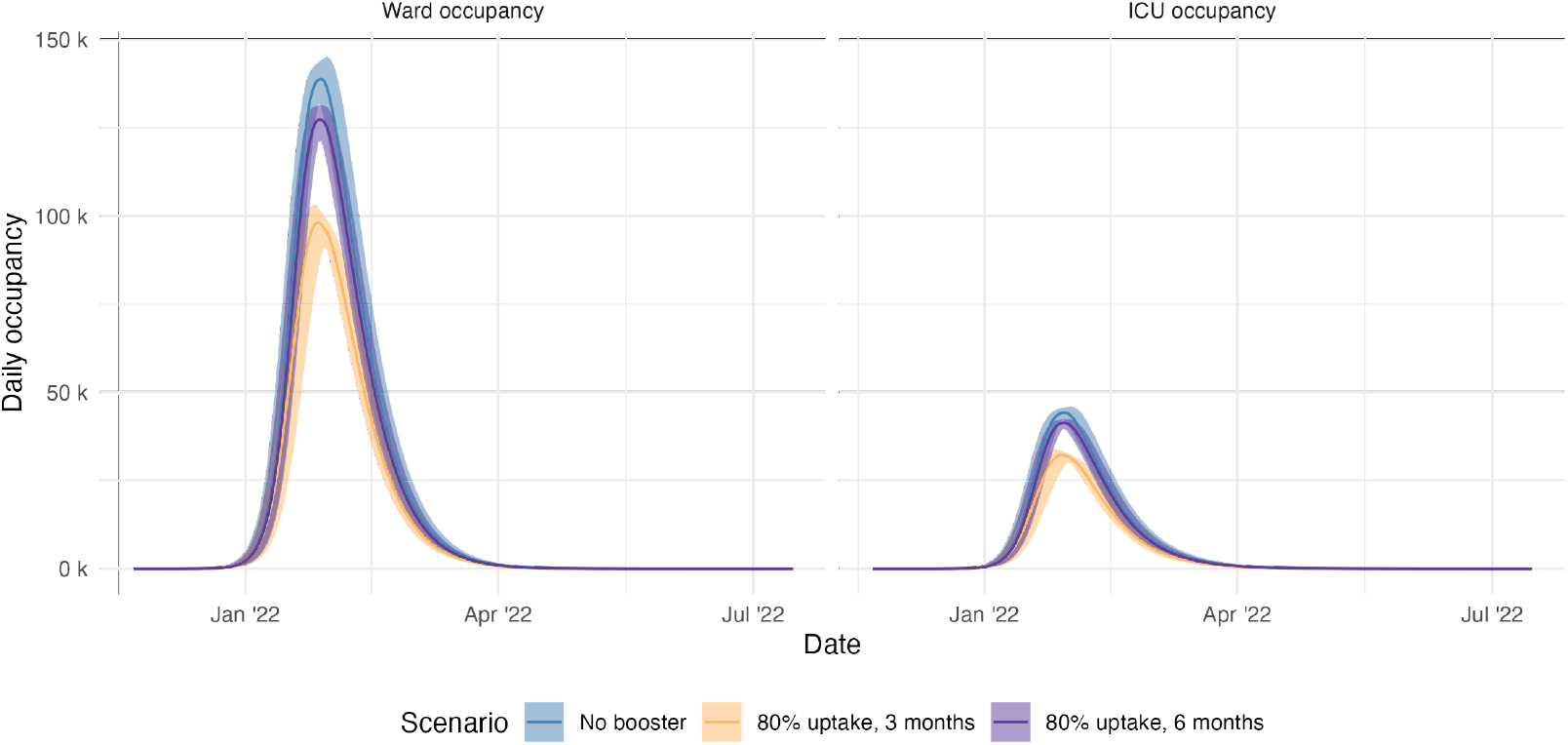
The ward and ICU occupancy levels for the projected Omicron wave, assuming the same severity as the delta strain.

**Fig 13.**
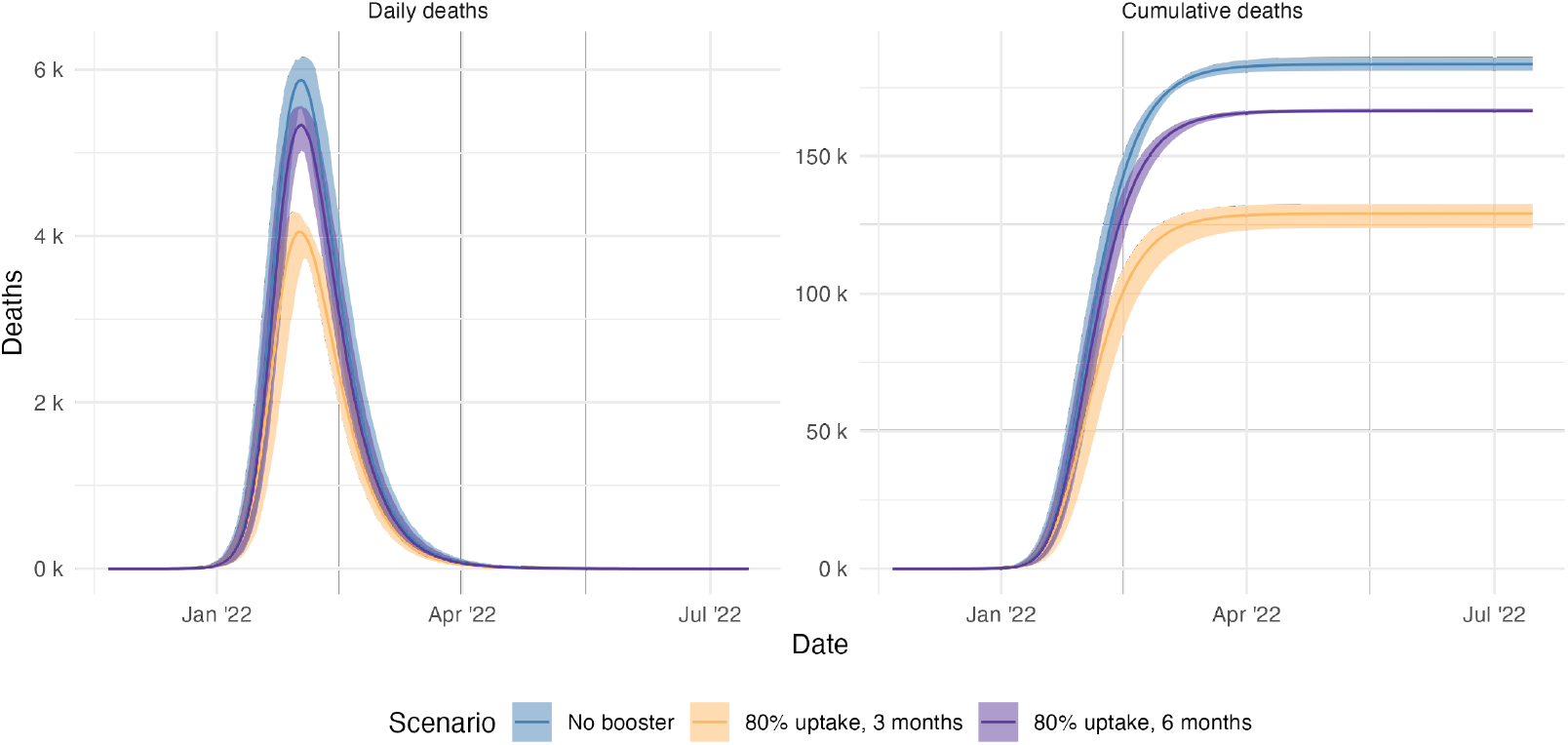
The daily and cumulative deaths for the projected Omicron wave.

#### 6.4.2 Protection against symptoms given infection

A typical study to measure vaccine efficacy against symptoms will be a test negative control, comparing those who were symptomatic and positive to those that were symptomatic and negative. Therefore, instead of measuring probability of symptoms given infection, the study is indirectly measuring the probability of symptoms and infection. In reality, the study would need to consider asymptomatic infections to get the desired probability of symptoms given infection.

After the study is finished, we can relate the probability of symptomatic infections given vaccine product, *p*(*S* ∩ *I*|*V*), to that of an unvaccinated individual through,

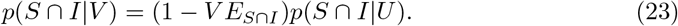

where *p*(*S* ∩ *I* |*U*) is the probability of symptomatic infection given you are unvaccinated.

We also know that

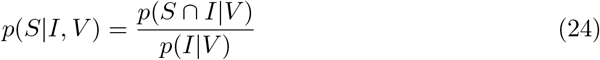

from the laws of probability. Therefore, we should be able to use the probability of infection given vaccination from another study to arrive at the correct probability, *p*(*S* | *I, V*).

Combining Equation 22, Equation 23 and Equation 24 we obtain,

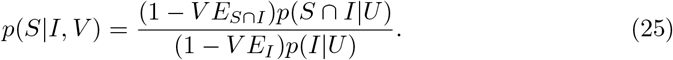

Note that,

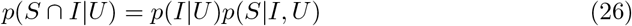

where *p*(*S*|*I, U*) is the probability of developing symptoms given infection in an unvaccinated individual. This is the baseline clinical fraction, which we denote as *q*_0_. Therefore,

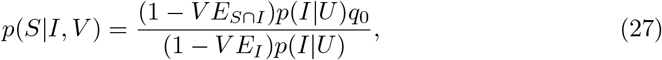

which simplifies to

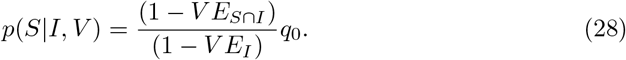

#### 6.4.3 Protection against onward transmission

It is typical to assume that asymptomatic individuals are less infectious when compared to their symptomatic counterpart. However, the measured vaccine efficacy against onward transmission does not account for the vaccine’s impact on reducing the symptomatic fraction. If the vaccine reduces the probability of developing symptoms, which in turn reduces the rate of onward transmission from the infected individual, than a naive implementation of vaccine efficacy against onward transmission will result in the “double counting” of vaccine efficacy. This double counting will result in increased overall efficacy of the vaccine.

The probability of transmission given that you are infected and vaccinated is given by

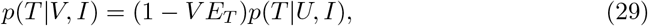

where *VE*_*T*_ is the vaccine efficacy for onwards transmission and *p*(*T* | *U, I*) is the probability of transmission given you are infectious and unvaccinated. We now assume that asymptomatic individuals are *α* times infectious as symptomatic individuals.

Therefore, we can expand either side of Equation 29 as

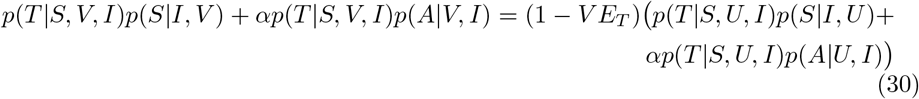

where *p*(*A*|*V, I*) and *p*(*A*|*U, I*) are the probability of being asymptomatic given infection if you are vaccinated or unvaccinated respectively. Defining *q* = *p*(*S*|*V, I*) (see Eq.(28)) and *q*_0_ = *p*(*S*|*U, I*) we obtain,

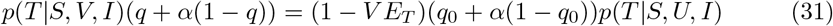

which can be simplified to

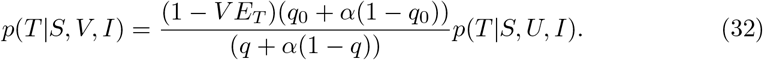

Note that if being asymptomatic does not reduce your infectivity (*α* = 1) then we do not have to correct *V E*_*T*_. On the other hand, if *α* = 0.5 (typical assumption),

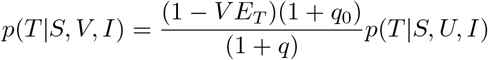

which is the expression that is implemented in our IBM to ensure we do not double count the effect of the vaccine.

### 6.5 Length of stay distributions

Length of stay in each model compartment were assumed to be Gamma distributed. Uncertainty was incorporated by sampling rate and shape parameters for each simulation from the posterior samples estimated from the Australian Delta wave [11], individual lengths of stay in each compartment were then sampled from the corresponding Gamma distributions. The mean lengths of stay in each compartment by age are given in Table 3.

**Table 3.** Mean length of stay in model compartments by age group.

| Age | [0, 40) | [40, 70) | 70+ |
| --- | --- | --- | --- |
| $I$ | 3.28 | 3.35 | 2.89 |
| $H_R$ | 3.61 | 5.79 | 12.33 |
| $H_D$ | 12.11 | 12.11 | 12.11 |
| $ICU_{pre}$ | 1.66 | 2.07 | 2.92 |
| $ICU_D$ | 17.94 | 17.94 | 17.94 |
| $ICU_{W_R}$ | 8.91 | 8.91 | 9.06 |
| $ICU_{W_D}$ | 8.91 | 8.91 | 9.06 |
| $W_R$ | 7.35 | 10.07 | 14.28 |
| $W_D$ | 5.67 | 5.67 | 5.67 |

**Table 4.** Transition probabilities by age for unvaccinated individuals with the wild-type strain.

| age | $p_{I\vert E}^w$ | $p_{H\vert I}^w$ | $p_{ICU\vert H}^w$ | $p_{H_D\vert ICU^c}^w$ | $p_{ICU_D\vert ICU}^w$ | $p_{W_D\vert ICU_D^c}^w$ |
| --- | --- | --- | --- | --- | --- | --- |
| [0, 5) | 0.28 | 0.029 | 0.058 | 0.018 | 0.189 | 0.032 |
| [5, 10) | 0.28 | 0.001 | 0.069 | 0.017 | 0.192 | 0.029 |
| [10, 15) | 0.2 | 0.005 | 0.081 | 0.016 | 0.195 | 0.027 |
| [15, 20) | 0.2 | 0.007 | 0.093 | 0.016 | 0.200 | 0.026 |
| [20, 25) | 0.26 | 0.020 | 0.106 | 0.017 | 0.208 | 0.026 |
| [25, 30) | 0.26 | 0.030 | 0.121 | 0.018 | 0.220 | 0.027 |
| [30, 35) | 0.33 | 0.032 | 0.137 | 0.021 | 0.237 | 0.028 |
| [35, 40) | 0.33 | 0.034 | 0.157 | 0.025 | 0.261 | 0.030 |
| [40, 45) | 0.4 | 0.038 | 0.181 | 0.034 | 0.299 | 0.033 |
| [45, 50) | 0.4 | 0.056 | 0.208 | 0.049 | 0.348 | 0.036 |
| [50, 55) | 0.49 | 0.104 | 0.229 | 0.072 | 0.405 | 0.041 |
| [55, 60) | 0.49 | 0.149 | 0.240 | 0.110 | 0.472 | 0.052 |
| [60, 65) | 0.63 | 0.185 | 0.233 | 0.162 | 0.540 | 0.074 |
| [65, 70) | 0.63 | 0.311 | 0.205 | 0.231 | 0.602 | 0.116 |
| [70, 75) | 0.69 | 0.479 | 0.155 | 0.311 | 0.649 | 0.184 |
| [75, 80) | 0.69 | 0.750 | 0.097 | 0.383 | 0.670 | 0.264 |
| 80+ | 0.69 | 0.655 | 0.026 | 0.460 | 0.615 | 0.350 |

### 6.6 Clinical probabilities given Delta infection in immune naive individuals

Transition probabilities for the wild-type strain in immune naive individuals from [10] and [21] are presented in Supplementary Table 4. We adjust these wildtype probabilities to account for the increased odds of severe outcomes for the Delta strain, these are adjusted again to account for the protection from past vaccination and infection (according to individual neutralisation titres).

The odds ratio of Delta strain compared to wild-type for hospitalisation (Δ_*h*_), ICU (Δ_*ICU*_) and death (Δ_*d*_) given infection are 2.08, 3.35, 2.33 respectively [22]. To apply these odds ratios we first combine marginal probabilities and then apply the odds ratio.

For example, we could multiply probability of symptoms 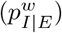 and probability of hospitalisation given symptoms 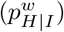 to get the probability of hospitalisation given infection 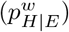 and apply the odds ratio of 2.08 to get the probability of hospitalisation given a Delta infection (*p*_*H*|*E*_). Let *OR*(*p, r*) denote the function that applied odds ratio *r* to probability *p*. We compute the following:

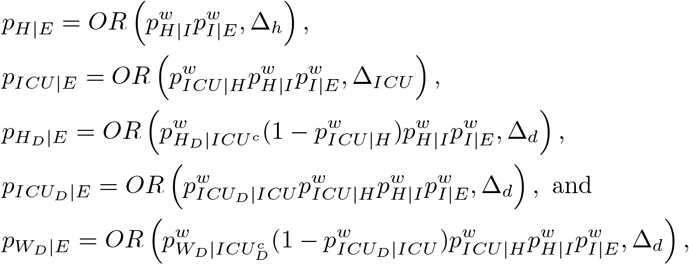

where

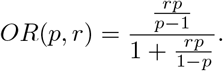

Note that we assume that the odds ratio relating to death given infection can be applied separately to all three death related components of the model. These baseline probabilities allow us to account for neutralisation titres and then marginalise the probabilities to obtain model transition probabilities.

### 6.7 Emergency Department Capacity Queue

Code for the clinical pathways model also allows for modelling of queues to the emergency department (ED). This was incorporated as it has been noted that limited ED consult capacity can cause a bottleneck that prevents admission to hospital [23]. However, this functionality was been omitted from the main text (by setting the ED queue to be infinite) so as to show pure hospital resource demand without considering capacity constraints.

The ED queue model is incorporated once it is determined that individual *i* requires hospital (via the random variable, *H*), and before the individual is admitted to hospital. If individual *i* requires hospitalisation they will present to the ED, where they may not be seen due to capacity limitations. ED consult capacity is modelled by admitting only the first *C*_*ED*_ presentations to ED each day. If individual *i* is not seen, they enter a state *ED*_*Q*_ they will present again to the ED with probability 1 − *p*_L|ED_ after *τ*_L|ED_ days sampled from

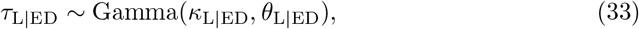

where *p*_L|ED_ is the probability that an individual does not present again to the ED and *κ*_L|ED_ and *θ*_L|ED_ are the shape and rate parameters of the gamma distribution respectively. For individuals that do not return to ED and are therefore not admitted to hospital, their age, neutralisation titre upon exposure and number of presentations to ED are recorded such that these can be used to understand possible excess mortality due to ED capacity limits. If individual *i* is admitted to hospital we determine what hospital pathway they will follow.

## Notes

### Competing Interest Statement

The authors have declared no competing interest.

### Funding Statement

This work was funded by the Australian Government Department of Health and Ageing Office of Health Protection. Additional support was provided by the National Health and Medical Research Council of Australia through its Centres of Research Excellence (SPECTRUM, GNT1170960).

